# Age-related differences in Rostral-Middle Locus Coeruleus Microstructure: A Critical Role in Cognitive Decline Revealed by Magnetic Resonance Relaxometry

**DOI:** 10.1101/2025.02.17.25322421

**Authors:** Jonghyun Bae, Zhaoyuan Gong, Caio Mazucanti, Murat Bilgel, John P. Laporte, Mary E. Faulkner, Alex Guo, Christopher M. Bergeron, Josephine M. Egan, Susan M. Resnick, Christopher E. Ramsden, Mustapha Bouhrara

## Abstract

**BACKGROUND:** The Locus Coeruleus (LC) is a critical brain region affected by neurodegenerative diseases and cognitive decline in aging. Despite its importance, in-vivo investigations of age-related LC degeneration and association with cognitive decline have been limited.

**METHOD:** We employed the Bayesian Monte-Carlo analysis of multicomponent driven equilibrium single pulse observation of T_1_ and T_2_ (BMC-mcDESPOT) method to estimate longitudinal (R_1_) and transverse (R_2_) relaxation rates in the LC of a diverse cohort of cognitively unimpaired individuals aged 22 to 94 years. BMC-mcDESPOT offers high spatial resolution and is effective for mapping detailed microstructural changes within the LC as reflected by R_1_ and R_2_. We examined age-related differences in LC microstructure, their associations with cognitive changes, and the spatial variation of these microstructural changes within the LC, exploring their distinctive contributions to cognitive decline.

**RESULTS:** Age was significantly associated with LC microstructural integrity, with advanced ages exhibiting lower R_2_ values. We also found that lower LC- R_1_ and LC-R_2_ are correlated with steeper decline longitudinally in multiple cognitive domains, including memory, verbal fluency, processing speed and executive function. Furthermore, we observed distinctive spatial variations between young and old participants, with R_1_ and R_2_ values in the rostral-middle regions being more strongly associated with cognitive changes as compared to the caudal region.

**CONCLUSIONS:** This study reveals age-related differences in LC integrity, which are associated with cognitive decline. LC relaxometry metrics (R_1_ and R_2_ values) may serve as sensitive biomarkers for detecting early alterations and age-related cerebral degeneration.

## 1. Background

The locus coeruleus (LC), a nucleus in the rostral pontine brainstem (14.5 mm in length and 2.5 mm in thickness on average) [1], plays a crucial role in the brain’s noradrenergic system. The LC’s noradrenergic neurons project to cortical and subcortical regions, modulating sleep and wake cycles [2], memory [3], and attention [4]. LC degeneration occurs in neurodegenerative diseases, such as Alzheimer’s disease (AD). Previous preclinical studies have linked LC impairments to amyloid pathology, neuroinflammation, and neurovascular dysfunction [5]. Moreover, LC degeneration is more strongly associated with tau pathology, with evidence suggesting that the first alterations in tau, such as phosphorylated tau accumulation, occur in the LC [6]. Additionally, a previous study found tau lesions in the LC in 94% of individuals by the age of 50, highlighting the prevalence of tau pathology [7]. Further, several studies [2, 8, 9] suggest spatially differentiated LC structure, where rostral LC neurons make projections to hippocampal regions and modulate cognitive functions, such as learning and memory, by releasing norepinephrine (NE) [10–12]. On the contrary, caudal regions of LC neurons innervate the cerebellum and spinal cord [13, 14]. Based on findings that highlight the crucial role of the LC in age-related cognitive decline [15, 16] and its implications in the progression to neurodegenerative diseases [17], it is essential to develop in vivo biomarkers that capture topographical changes in LC integrity. Although several MRI studies have attempted to assess LC integrity, these methods often lack specificity and are less quantitative. We therefore aim to fill in this deficit and apply state-of-the-art MR imaging of the LC.

Previous MRI studies have identified differences in LC structure using T_1_-weighted images obtained with turbo spin echo sequences [18]. The neuromelanin-rich LC neurons exhibit a T_1_-shortening effect, appearing hyperintense [19]. Most studies assess LC integrity by calculating the contrast ratio (CR) between LC signal intensity and a reference region [8, 20–23], while a few have attempted to measure volumetric changes [24]. While a previous study [25] reported the association of reduced CR with indicators of AD pathology, other studies present conflicting results [26, 27]. Similarly, studies on LC changes during normal aging have yielded inconsistent results [28, 29]. Moreover, these MRI studies mostly employ 2D imaging approach with thick slices of ∼3mm, potentially hindering accurate assessment in topographical changes of LC structure. Despite their potential for monitoring aging processes and pathological development, there is an unmet need for quantitative metrics to assess LC microstructural integrity and its impact on cognitive functioning. Microstructural changes have been shown to precede, by decades, macrostructural changes [30]. Therefore, besides establishing imaging biomarkers for early detection of abnormal changes in LC microstructure, such investigations are critical to decipher the mechanisms underlying structural changes in the LC and to differentiate those due to normal from abnormal aging. The paucity of such studies is perhaps due to the small size of LC. Therefore, it is essential to use high spatial resolution imaging to minimize partial volume effects.

Despite its’ effectiveness in probing tissue microstructure and composition of the brain [31], MR relaxometry has not yet been tested for evaluating the structural integrity of the LC. As demonstrated in previous MRI studies, neuromelanin – a by-product of catecholamines – is paramagnetic and shortens longitudinal relaxation time (T_1_ = 1/R_1_). Therefore, a reduction in R_1_ may indicate alterations in the proper functioning of the noradrenergic system. Additionally, transverse relaxation time (T_2_ = 1/R_2_) derived from MR relaxometry may reflect microstructural integrity, such as cellular density, as reflected by a high correlation with apparent diffusion coefficient (ADC) [32]. To evaluate the structural integrity of the LC, we employed the Bayesian Monte-Carlo analysis of multicomponent driven equilibrium single pulse observation of T_1_ and T_2_ (BMC-mcDESPOT) method [33] for high spatial resolution mapping of relaxation rates (R_1_, R_2_) [33, 34]. This quantitative MRI (qMRI) relaxometry technique has been adopted in several clinical investigations of central nervous system maturation and degeneration [35–38].

Our study cohort consists of 113 cognitively unimpaired participants spanning a well-characterized age range of 22 to 94 years. BMC-mcDESPOT has been used extensively to investigate the patterns of cerebrum and brainstem myelination, composition and microstructural changes with aging and dementias [39, 40]. Our main goals are; *i*) to investigate age-related differences in LC microstructure and disruptions in adrenergic system, and, *ii*) to study the association of differences in LC microstructure and alterations to adrenergic system with changes in cognition. We herein test the hypotheses that; *i*) LC’s microstructural integrity is lower at advanced ages, and, *ii*) lower microstructural integrity of LC is associated with lower memory retention and attention, and with their steeper declines over time. This study aims to deepen our understanding of the relationship between LC’s structural integrity with age and cognition. This knowledge may allow for developing imaging biomarkers sensitive to early changes in LC to help monitoring targeted interventions in the brain.

## 2. Methods

### 2.1. Participants

The study sample comprised participants drawn from two National Institute on Aging / National Institutes of Health Intramural Research Program (NIA/NIH IRP)-approved studies: the Baltimore Longitudinal Study of Aging (BLSA; Protocol #03AG0325) [41] and the Genetic and Epigenetic Signatures of Translational Aging Laboratory Testing (GESTALT; Protocol #15AG0063). Both studies adhered to identical inclusion and exclusion criteria, with a shared objective of assessing multiple aging-related biomarkers. Prior to enrollment in our MRI protocol, stringent eligibility criteria were applied, ensuring the absence of central nervous system diseases (e.g., dementia, stroke, bipolar illness, epilepsy), cardiac disease, pulmonary disease, and metastatic cancer. Participants with metallic implants or significant neurological or medical disorders were excluded from the MRI cohort. Participants were determined to be cognitively normal if they had ≤ 3 errors on the Blessed Information-Memory-Concentration (BIMC) Test [42], and where available, a Clinical Dementia Rating (CDR) [43] of zero. If participants had > 3 errors on the BIMC or a CDR > 0, their clinical and neuropsychological data were thoroughly reviewed as part of consensus case conferencing. Visits where participants were found to have cognitive impairment or dementia were excluded from these analyses.

### 2.2 Neuropsychological assessment

Cognitive domain scores were obtained for memory (California Verbal Learning Test [44] immediate and long-delay free recall), attention (Trail Making Test Part A (TMT A) [45] and Digit Span [46] Forward), executive function (Trail Making Test Part B and Digit Span Backward), verbal fluency (Category [47] and Letter Fluency [48]) and processing speed (Digital Symbol Substitution Test (DSST) [49]). Each test score was first converted to a z-score using the baseline mean and standard deviation. When multiple tests were administered within a cognitive domain, these z-score were averaged to compute the domain score. For processing speed, we adopted a composite score as suggested before [50], and combined results from TMT A and DSST. We note that individual scores for Trail making Test Parts A and B were log-transformed and negated prior to computing z-scores, so that all high z-scores reflect shorter completion times. Figure 1 demonstrates the distribution of test scores against participant’s age for each cognitive domain. Most participants (∼63%) underwent cognitive evaluation for memory at least twice at or before the MRI scan. For other cognitive domains, ∼50% of participants had at least two cognitive evaluations before or at the time of MRI scan.

**Figure 1.**
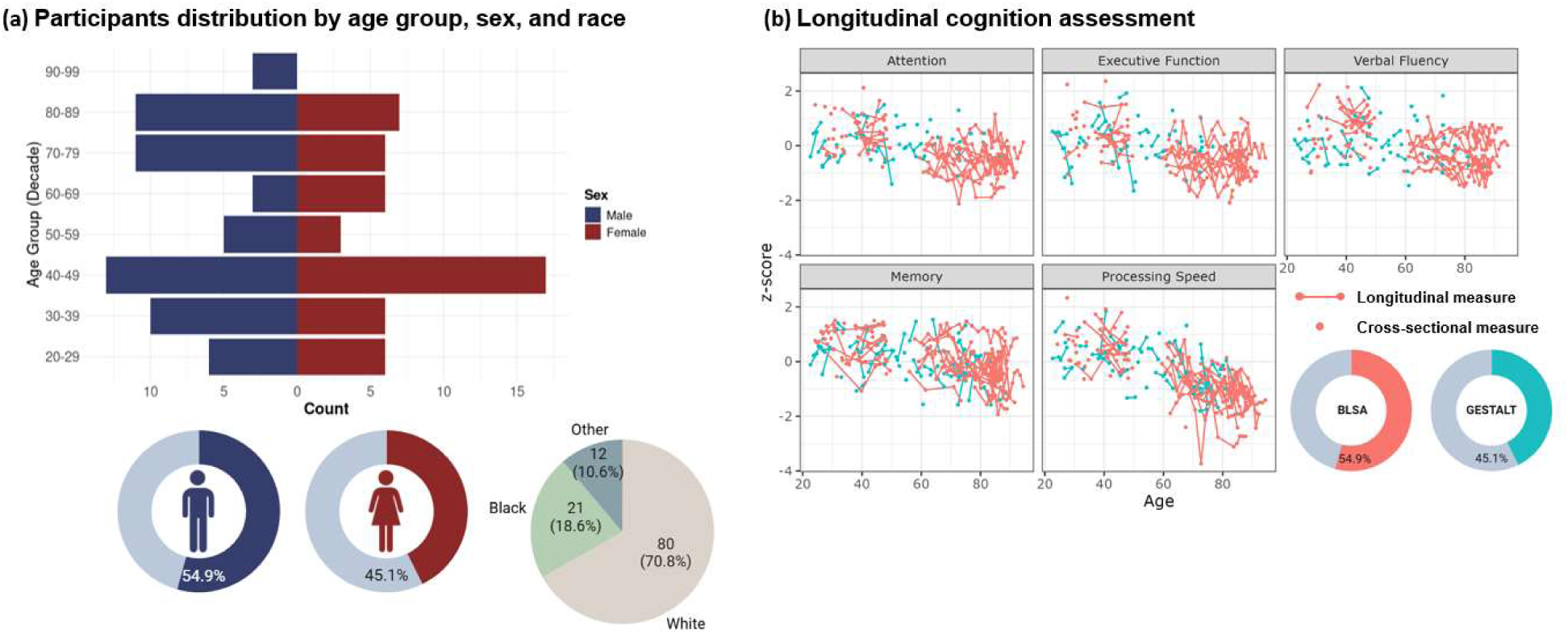
(a)Participants distribution by age group, sed and race. Our study included participants aged 22 and 94 years, with a well-balanced distribution between sexes and a fair representation of both young and old age groups. (b)Longitudinal cognitive trajectories across multiple domains. This graphical representation displays z-scored measures for various cognitive domains, with each panel showcasing a different domain. Longitudinal cognitive data for each participant are represented by a dot and connected lines, illustrating individual trajectories over time. The cohorts are distinguished by color, with red indicating participants from the Baltimore Longitudinal Study of Aging (BLSA) and blue representing those from the Genetic and Epigenetic Signatures of Translational Aging Laboratory Testing (GESTALT) study.

### 2.3. MRI acquisition

All MR scans were acquired on a 3T whole-body Philips MRI system (Achieva, Best, The Netherlands) using the internal quadrature body coil for radio-frequency (RF) transmission and an eight-channel phased-array head coil for reception. Each participant underwent our BMC-mcDESPOT protocol [33], consisting of 3D spoiled gradient echo (SPGR) sequences (TE/TR = 1.48/5 ms) and balanced steady state free precession (bSSFP) sequences (TE/TR = 2.8/5.9 ms) with RF excitation pulse phase increment of 0 and 𝜋 to account for off-resonance artifacts [51]. All three sequences were acquired with multiple flip angles: [2, 4, 6, 8, 10, 12, 14, 16, 17, 20]° for SPGR and [2, 4, 7, 11, 16, 24, 32, 40, 50, 60] ° for bSSFP sequences. All images were acquired with acquisition matrix of 150 × 130 × 94 and an isotropic voxel size of 1.6 mm, reconstructed to a 1 mm isotropic resolution. To correct the RF inhomogeneity, 𝐵_1_, the double-angle method [52] was implemented with two fast spin-echo images acquired with FAs of 45° and 90° (TE/TR = 102/3000 ms, spatial resolution = 2.6 × 2.6 × 4 𝑚𝑚). Estimated low-resolution 𝐵_1_ field was further interpolated to align with the same spatial resolution of the SPGR and bSSFP images.

### 2.4 Data processing and region segmentation

For each participant, a whole-brain R_1_ map was generated from the SPGR and DAM datasets using DESPOT1 and assumes a single component, and a whole-brain R_2_ map was generated from the bSSFP and DAM datasets using DESPOT2 assuming a single component [53]. The averaged SPGR image for each case was registered to the Montreal Neurological Institute (MNI) template (MNI-ICBM 152 linear space, 0.5 mm resolution), using Advanced Normalization Tools [54]. Using the resulting transformation matrix, individual R_1_ and R_2_ were then warped to the template space. For LC localization, we adopted a meta-mask provided by Dahl et al. [55], which combines previously reported LC masks to enhance the identification of small LC structures. Using the meta-mask, we calculated the mean values of R_1_ and R_2_ across the entire LC structure for each participant. To achieve topographical assessment of LC, we computed slice-averaged R_1_ and R_2_ values. Then, we adopted a similar approach to a previous study [15] and identified rostral-middle (29 – 64^th^ LC rostro-caudal percentile) and caudal (64 – 100^th^ LC rostro-caudal percentile) regions of the LC. Additionally, we evaluated the mean values of R_1_ and R_2_ for both the rostral-middle LC and caudal regions of the LC.

### 2.5 Statistical analysis

#### Age-related differences in the LC integrity

We first examined differences in LC microstructure or myelination with aging. Each of the LC-qMRI measures (*i.e.*, R_1_ and R_2_) was correlated with age while controlling for the effects of sex, years of education (EDY) and race using the following multiple linear regression model:

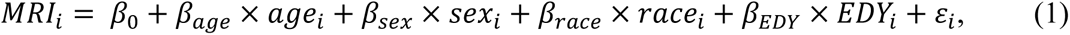

where 𝑀𝑅𝐼_i_ is R_1_ or R_2_, 𝑎𝑔𝑒_i_ is age of the *i^th^* participant at the time of MRI, and 𝜀_i_ is the residual.

We also examined for non-linear trends in changes to LC integrity, as suggested by other studies [56]. The non-linear model accounting for potential inverted U-shaped trend can be described as follows:

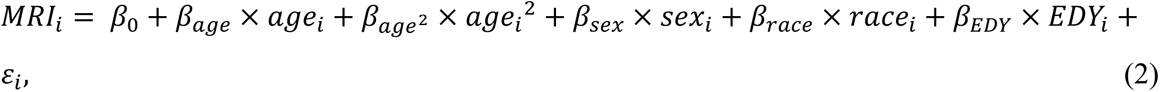

where 𝑎𝑔𝑒_i_^2^ is age-squared of the *i^th^* participant at the time of MRI, and 𝜀_i_ is the residual. Age was mean centered to avoid collinearity with the age^2^ term.

#### Association between LC integrity and cognition

Then we conducted a cross-sectional analysis of the association between each of the LC-qMRI measures and each of the five cognitive domains. The dependent variable was each of the z-scored cognitive domain at the time of MRI while the independent variables were age at the time of MRI, sex, race, EDY and LC-qMRI using the following multiple linear regression model:

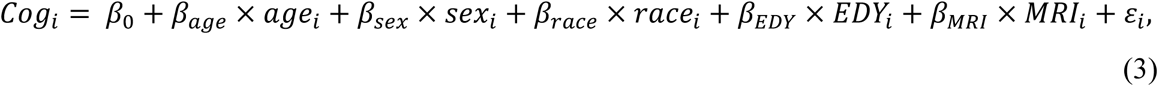

where 𝐶𝑜𝑔_i_ is the cognitive domain score at the time of MRI.

#### Association with changes in cognition

We investigated the association between each of the LC-qMRI measures (*i.e.*, R_1_ and R_2_) and longitudinal changes in cognition. We employed separate linear mixed-effects models for each cognitive domain. The longitudinal cognitive scores were considered as the dependent variables while age at MRI, sex, race, EDY, LC-qMRI measures, and LC-qMRI measure × time interaction were the independent variables. The linear mixed effect model for the longitudinal analysis is given by:

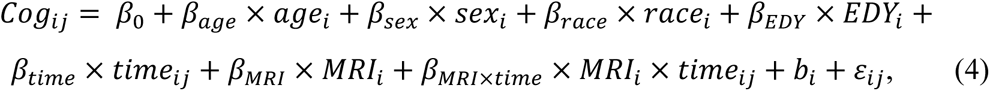

where 𝐶𝑜𝑔_ij_ is the cognitive score of subjects 𝑖 at time 𝑗, 𝑡𝑖𝑚𝑒_ij_is the time from MRI of the *i^th^* participant at time point 𝑗, and 𝑏_i_ is the corresponding random intercept. To facilitate interpretation, LC-qMRI values were mean-centered. We note that the main parameter of interest in this analysis is 𝛽_MRI×time_, reflecting the expectation of the difference in the annual change in cognition per unit difference in R_1_ or R_2_. Each metrics were corrected for false discovery rate (FDR), using the Benjamini-Hochberg procedure [57].

#### Topographical differences in LC integrity between rostral-middle and caudal regions across aging

After computing the mean values of qMRI metrics for each slice, we compared the slice profiles between the young (< 60 years) and old groups. A two-sample t-test was performed at each slice to assess age-related topographical differences in qMRI metrics. We also examined changes in qMRI metrics across the age spectrum, stratified by the rostral-middle and caudal regions of the LC. We conducted a paired t-test to assess whether age-related changes in qMRI metrics vary across different regions of the LC.

#### Association between LC rostro-caudal integrity and changes in cognition

We tested the association between qMRI measures in sub-regions of the LC (i.e. rostral-middle and caudal) and longitudinal changes in cognition. We applied similar models as described above, while separate models were established for rostral-middle and caudal regions. The linear mixed effect model for this analysis is provided by:

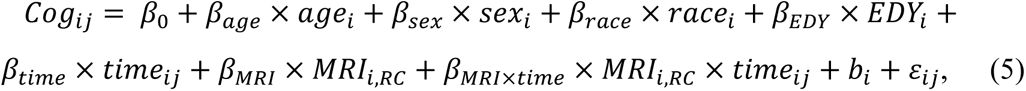

where 𝐶𝑜𝑔_ij_ is the cognitive score of subjects 𝑖 at time 𝑗, 𝑡𝑖𝑚𝑒_ij_ is the time from MRI of the *i^th^* participant at time point 𝑗, 𝑅𝐶 regional mean of qMRI metrics in either rostral-middle or caudal LC regions and 𝑏_i_ is the corresponding random intercept. Again, our primary focus was the interaction term between time and qMRI metrics in sub-regions of the LC, examining how changes in these metrics contribute to annual changes in cognition. The resulting 𝑝-values were corrected for multiple comparisons in the same manner.

## 3. Results

Table 1 shows demographic information of our study cohort. Our cohort consisted of 113 subjects spanning the age range of 22 to 94 years. The participants were well-balanced in terms of sex distribution (55% male), and there were no significant differences in age between males and females (*p* > 0.1). Racial distribution was predominantly White (71%) followed by Black (19%). Participants had an average of 16 years of education. As shown in Figure 1, 93% of the participants had MRI scans within 2 years of the last neuropsychological assessment with only one participant undergoing the MRI scan 3 years after the last neuropsychological testing.

**Table 1.** Participant demographic information.

| **Characteristics** | **Cognitively normal participants**<br>**(n = 113)** |
| --- | --- |
| **Age at MRI scan, mean (SD)** | 56.2 (20.6) |
| **Sex, Men (%)** | 62 (54.9%) |
| **Race, White (%)** | 80 (70.8%) |
| **Race, Black (%)** | 21 (18.6%) |
| **Education (yrs), mean (SD)** | 16.2 (2.7) |
| **Number of cognitive assess.,**<br>**mean (SD)** | 2.78 (3.0) |
| **Duration of cognitive assess.,**<br>**median (mean, SD)** | 2.1 (4.3, 6.4) |

Figure 2a illustrates the identification of LC structure using the meta-mask, overlaid on MNI template. Figure 2b shows quantitative results for the LC-qMRI (*i.e.*, R_1_, R_2_, and MWF) values derived from all participants as a function of age. Although LC-R_1_ did not exhibit any age-related differences, LC-R_2_ showed significant negative associations with age (𝑝_corr_ < 0.05). None of qMRI metrics exhibited quadratic trends with age. Although the cross-sectional analysis of associations between LC-qMRI and cognition revealed no significant correlations after FDR corrections, our longitudinal analysis indicates that lower LC-R_1_ and LC-R_2_ values are associated with steeper declines in memory and verbal fluency. Lower LC-R_1_ and LC-R_2_ values, corresponding to the value at the 25^th^ percentile as indicated by the red lines in Figure 3a, were significantly associated with steeper longitudinal decline in memory and verbal fluency (𝑝_corr_< 0.05).

**Figure 2.**
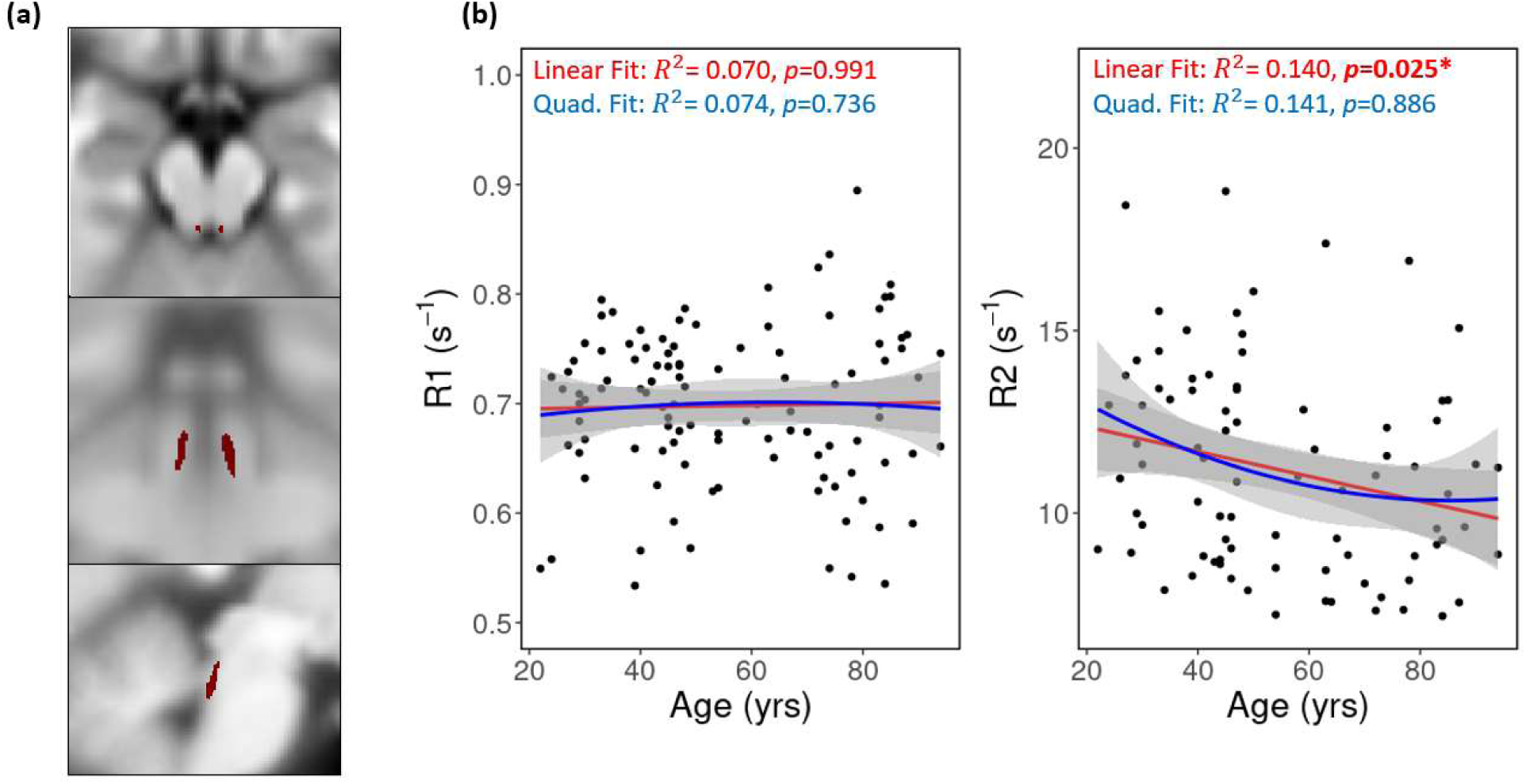
(a) The Locus Coeruleus (LC) meta-mask used in this study is overlaid on Montreal Neurological Institute (MNI) template (MNI-ICBM 152 linear space, 0.5 mm resolution) and presented in axial (top), coronal (middle) and sagittal (bottom) view. (b) Example of associations between LC quantitative MRI (LC-qMRI) metrics and age, adjusted for sex, race, and years of education. Both linear (red) and quadratic (blue) models were tested. Significant negative correlations were observed between age and LC-R_2_.

**Figure 3.**
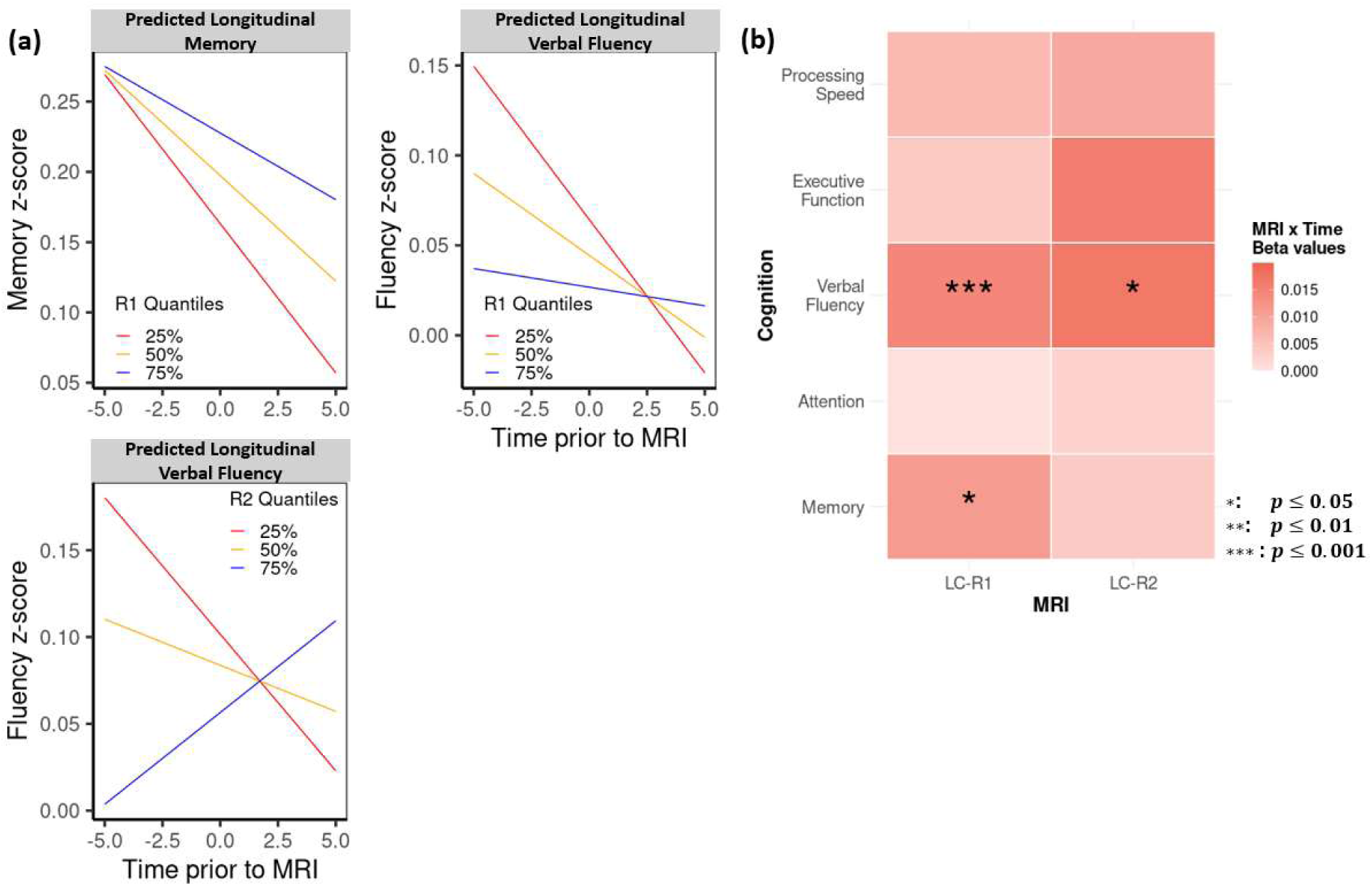
(a) Predicted longitudinal cognitive trajectories estimated from linear mixed-effects regression models at different quantiles of Locus Coeruleus quantitative MRI (LC-qMRI) metrics (red, orange, and blue lines represent the 25^th^, 50^th^, and 75^th^ percentiles, respectively). The results show significant differences in the rate of cognitive decline across various domains, including memory and verbal fluency for LC-R_1_. Additionally, LC-R_2_ showed significant associations with longitudinal changes in verbal fluency. These findings suggest that individual differences in LC-qMRI metrics are linked to distinct patterns of cognitive aging. (b) A heatmap illustrating the resulting coefficient for MRI×Time interaction terms from the linear mixed effects models across cognitive domains. All MRI measurements (i.e. LC-R_1_ and LC-R_2_) were mean-centered and resulting p-values were adjusted for False Discovery Rate correction.

Figure 4a shows the segmentation between the rostral-middle (29 – 64^th^ LC rostro-caudal percentile) and caudal regions (64 –100^th^ LC rostro-caudal percentile). Figure 4b, c displays the rostro-caudal profiles of LC-R_1_ and LC-R_2_ between young and old groups. These topographical profiles indicate higher LC-R_1_ and LC-R_2_ values in the rostral-middle regions compared to the caudal regions, except for LC-R_1_ in old group. Additionally, young groups exhibited higher LC- R_1_ and LC-R_2_ values than the old groups, except for LC-R_1_ in caudal regions. Only LC-R_2_ profile showed significant differences between two age groups. Figure 4d, e illustrates the qMRI metrics in both the rostral-middle and caudal LC as a function of age. The rostral-middle LC-R_1_ peaks around the age of 60 and gradually declines with age, while the caudal LC-R_1_ continues to increase following a relative plateau in middle age (Figure 4d). In contrast, LC-R_2_ displays similar age-related trends, showing a gradual decrease starting around the age of 50 (Figure 4e). Our paired t-test revealed significant differences between the rostral-middle and caudal regions for LC-R_2._

**Figure 4.**
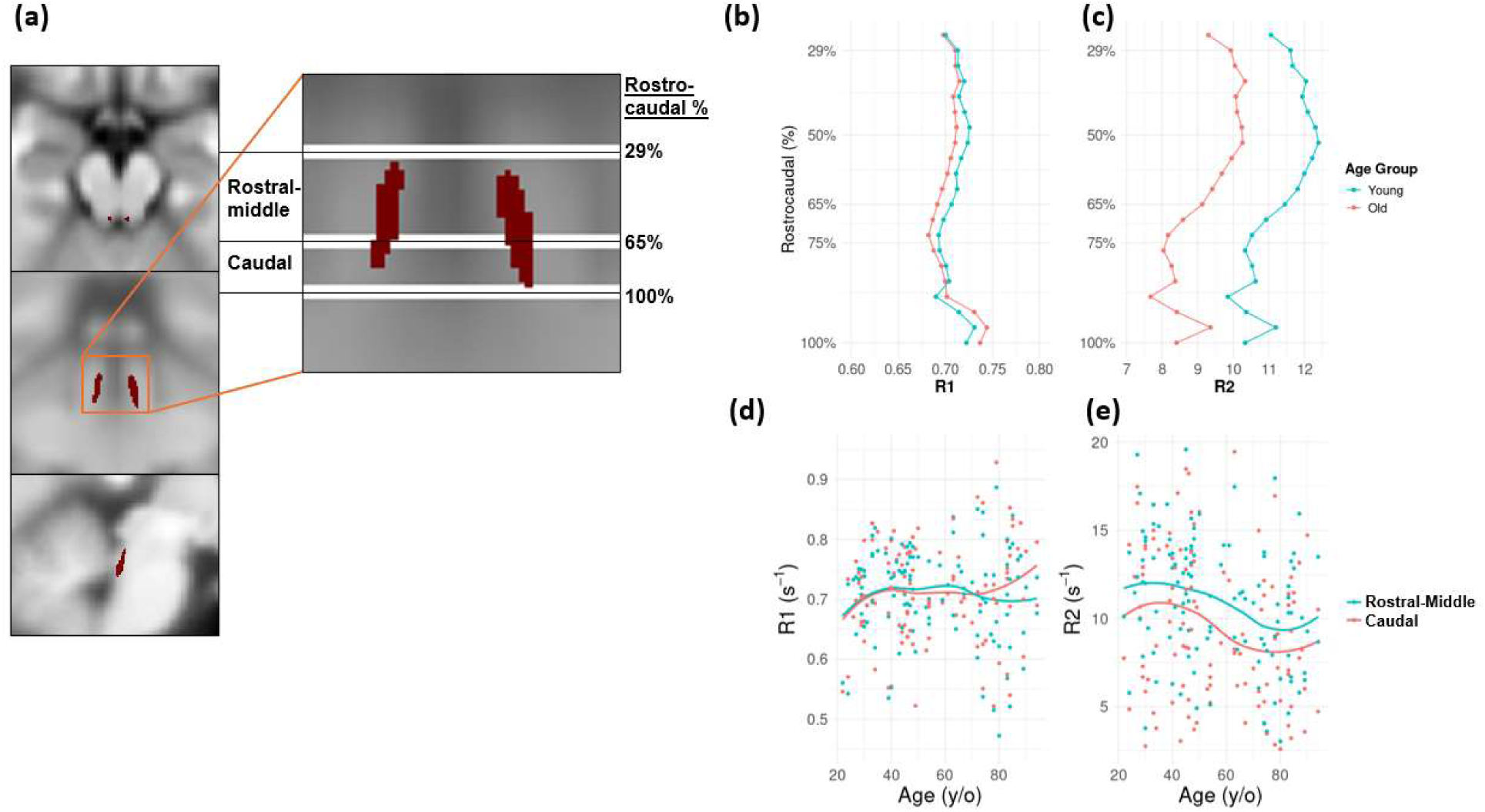
(a) Coronal view of LC structure for rostro-caudal identification. Adopted from a previous study [15], the rostral-middle (29 – 64% rostro-caudal percentile) and caudal (64-100^%^ rostro-caudal percentile) regions of the LC are delineated. (b-c) Rostro-caudal profiles of (b) LC-R_1_ and (c) LC-R_2_ between young (<60 years old) and old groups. The profiles revealed age-related reduction in both LC-R_1_ and LC-R_2_ values, except R_1_ values in the caudal LC region (d-e) Mean R_1_ and R_2_ values in the rostral-middle and caudal regions of the LC across age. (d) The rostral-middle R_1_ values peak at the age of 60 and subsequently decline, while the caudal R_1_ values continue to rise with age. (e) R_2_ values exhibit a similar trend in both regions, beginning to decline after the age of 50.

Finally, Figure 5 demonstrates how longitudinal changes in cognitions is associated with qMRI metrics in both the rostral-middle (Figure 5 a-c) and the caudal regions of the LC (Figure 5 d,e). Our results suggest that individuals with lower LC-R_1_ experience steeper declines in memory and verbal fluency and those with lower LC-R_2_ show more pronounced declines in executive function (𝑝_corr_< 0.05). To our surprise, we also found that both LC-R_1_ and LC-R_2_ in the caudal regions of the LC are significantly associated with greater declines in verbal fluency (𝑝_corr_ < 0.05).

**Figure 5.**
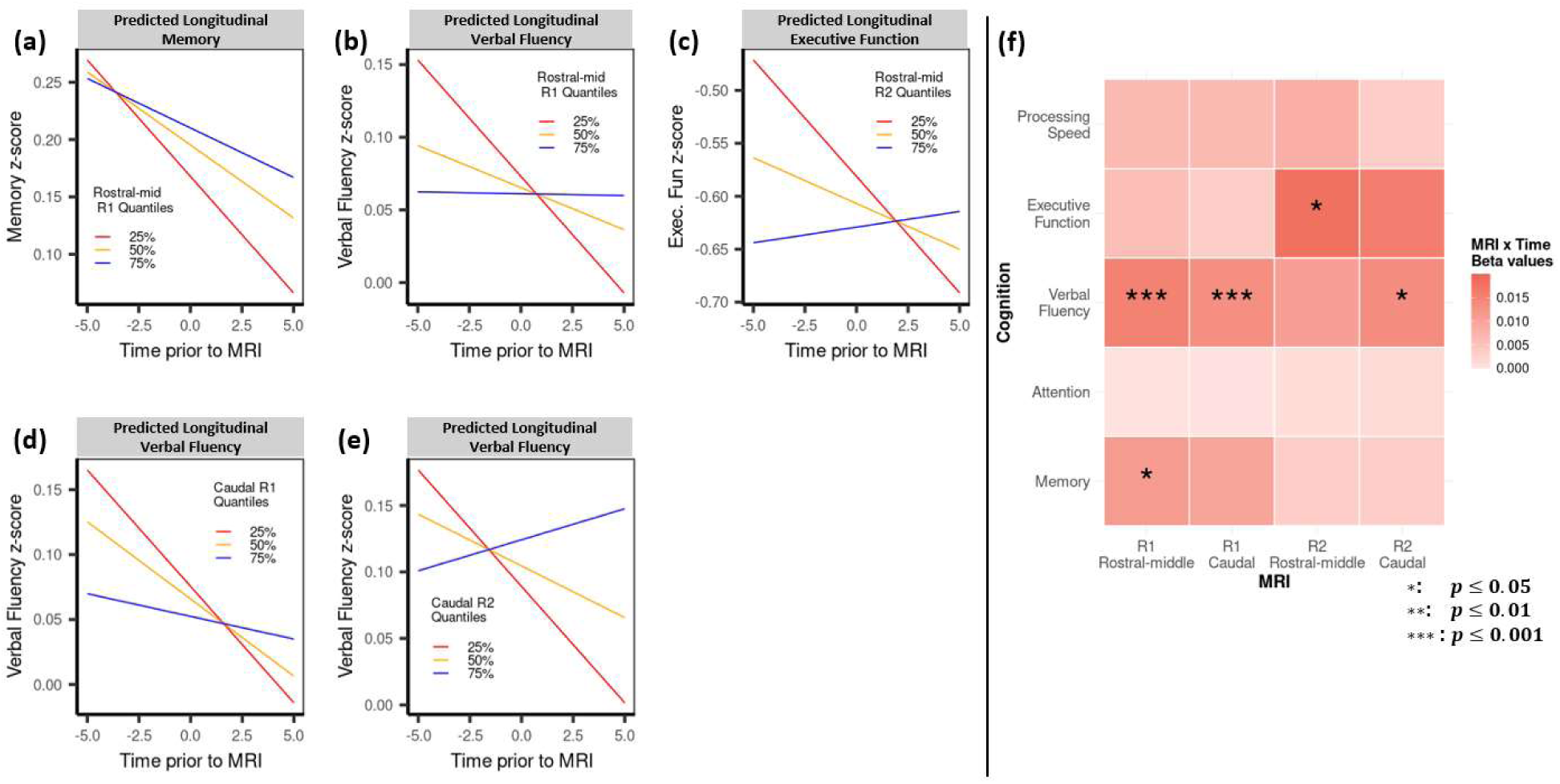
Predicted longitudinal cognitive trajectories estimated from linear mixed-effects regression models at different quantiles of qMRI metrics in subregions of the LC. (a-c) The results indicate that lower qMRI metrics in the rostral-middle LC are associated with steeper decline in multiple cognitive domains, such as memory, verbal fluency and executive function. (d-e) Lower R_1_ and R_2_ values in the caudal region are correlated with faster decline in verbal fluency. (f) A heatmap illustrating the resulting coefficient for MRI×Time interaction terms from the linear mixed effects models across cognitive domains. All MRI measurements (i.e. R_1_-Rostral-middle/Caudal and R_2_-Rostral-middle/Caudal) were mean-centered and resulting p-values were adjusted for False Discovery Rate correction.

## 4. Discussion

The LC contains over 50% of the brain’s noradrenergic neurons, serving as a central hub with projections to various brain regions, including the cortex, hippocampus, and thalamic nuclei [58]. NE release from the axons of LC neurons, particularly in rostral regions, stimulates hippocampal and cortical neurons, suggesting a crucial role for the LC in regulating cognitive functions, particularly memory and learning [59]. This mechanism has been partially observed in animal studies where transgenic rodents with tau pathology exhibited significant neuronal loss, memory loss, and learning deficiencies [60]. Similarly, post-mortem human brain studies found reduced LC volume associated with advanced Braak stages [61] and increased hyperphosphorylated tau levels in the LC during early AD pathology [62]. Furthermore, a significant progressive reduction in LC neurons was observed in Mild Cognitive Impairment (MCI) and AD patients compared to healthy subjects, which correlated with cognitive decline [63]. While these pre-clinical and ex-vivo studies suggest a close link between LC neuronal degeneration and the pathogenesis of neurodegenerative diseases, in-vivo MR imaging studies have yielded conflicting results. Some studies have found reduced LC-MR signal intensity in patients with neurodegenerative diseases, such as Parkinson’s disease [64], AD [26], and with major depression [65]. However, other studies have reported inconsistent findings, including no notable differences in LC signal between AD patients and healthy controls [66]. Additionally, age-related changes in LC MR signal further complicate the interpretation. For instance, Olivieri et al. [67] observed significant LC signal reduction in both AD and MCI groups compared to healthy controls, while Betts et al. [68] only observed such a change in the AD group, but not in the MCI group. These discrepancies call into question the sensitivity of the imaging measures used in these studies. Indeed, the commonly used contrast ratio measure may be biased by physiological changes in the reference tissue, which can subsequently bias the LC signal [20]. Furthermore, methodological differences in LC signal estimation, such as computing CR of LC-signal averaged from both left and right [68], using raw signal intensity [67] or localizing the signal to a specific location of LC [69], can also lead to inconsistencies. Additionally, the acquired signal intensity can substantially vary depending on the types of MR sequence, sequence parameters (e.g., flip angle, echo time, repetition time), and hardware (e.g., coil geometry, signal amplifier) [70], preventing direct measurement or comparison among subjects or across studies. Moreover, previous studies often used 2D imaging techniques with thick slice profiles. While thicker slices enhance signal-to-noise ratio, they may hinder accurate identification of the rostro-caudal LC. Further, variations in slice thickness could introduce inconsistencies in the computed LC-CR, further complicating the interpretation of the results. These limitations in using CR as a quantitative metric for LC integrity impair the accurate assessment of neuronal degeneration in the LC and reduce the sensitivity of in vivo, image-derived measurements.

In comparison, qMRI techniques provide imaging biomarkers that are sensitive to tissue microstructures and composition, enabling the assessment of subtle changes in brain tissue. Notably, diffusion tensor imaging (DTI) has been employed to quantitatively evaluate the microstructure of the LC. A post-mortem DTI study revealed increased fractional anisotropy (FA) in the brains of individuals with AD, suggesting a loss of noradrenergic cells and fibers, which is consistent with the neuropathological hallmarks of AD [71]. Additionally, Quanttrini et al. [72] demonstrated increased diffusivity along the LC-transentorhinal cortical pathway, a finding consistent with a stage subsequent to the initial spread of tau pathology in the LC [6]. Furthermore, the sensitivity of qMRI revealed microstructural changes in the LC during normal aging characterized by reduced mean and radial diffusivities and increased FA in older compared to younger individuals [73]. These studies also showed that these changes are significantly correlated with cognitive decline. However, diffusion imaging protocols are typically limited by low spatial resolution, as used in the aforementioned studies. These low-resolution images may hinder the accurate identification of microstructural changes in small structures, such as the LC. Alternatively, MR relaxometry, particularly BMC-mcDESPOT used in our study, provides whole-brain, isotropic high-resolution mapping of relaxation rates, which can serve as a proxy to infer tissue microstructures and composition. In addition, these metrics are less dependent on methodology or hardware compared to signal intensity-based features and have been extensively applied to various neuropathologies [74, 75].

Both longitudinal (R_1_) and transverse (R_2_) relaxation rates may be sensitive to changes in the tissue microstructural environment of the LC, and yet they may be influenced by different underlying mechanisms. Since T_1_-shortening neuromelanin is a by-product of NE synthesis, a reduction in R_1_ (increased T_1_) may indicate decreased NE production activity. In contrast, a reduction in R_2_ may indicate decreased cellular density, as reflected high correlation with ADC [32]. Based on previous findings from diffusion studies [73] in the LC, we conjecture that the age-related decline in R_2_ may reflect the reduction in adrenergic cells and fibers. Notably, we didn’t observe an age-related decline in LC- R_1_. Interestingly, LC- R_1_ was more sensitive towards changes in cognition, impacting memory, executive function and verbal fluency. These results suggest that decreased activity in noradrenergic system, as indicated by reduced LC- R_1_, impairs cognitive function. Indeed, LC neurons play extensive roles in hippocampal memory processing, which is involved in both memory formation and retention [76]. These neurons innervate three primary regions in the brain that establish functional connectivity to hippocampus: the basolateral amygdala for memory formation [77], ventral tegmental area for dopaminergic projections to hippocampus [78], and the prefrontal cortex for memory encoding and retrieval [79]. Furthermore, our subregion analysis revealed that the R_1_ reduction in the rostral-middle LC is specifically correlated with memory declines, but not with the caudal region. This is consistent with numerous studies [2, 9, 14] that identify the rostral-middle regions as the primary area influencing memory performance. Additionally, we observed our LC-R_1_ follows precisely the same age-related trajectory as reported in previous studies. For instance, neuromelanin accumulations in the rostral LC peaks around the age of 60 [80] and subsequently declines thereafter [81, 82]. The LC-R_1_ trend in Figure 4d exhibits nearly identical trends that directly align with these findings. In contrast, the caudal LC region exhibits continuous neuromelanin accumulation across the life span [81, 82], which is reflected as increased R_1_ (shortening T_1_) in our study (Figure 4d). Our observations in younger adults also align with a previous study, suggesting minimal differences between the rostral and caudal regions, likely due to the fact that many LC neurons have not yet accumulated neuromelanin [9]. Thus, our R_1_ metrics offer an accurate assessment of LC neuronal activity, which is crucial for maintaining cognitive function. We also observed an age-related decline of LC- R_2_ in both regions, but significantly higher R_2_ values in the rostral-middle, compared to the caudal region. There are several studies suggesting LC neuronal cell loss in healthy aging and in different neuropathology [9, 55, 83]. While previous studies indicate that this cell loss is specific to the rostral region, to our surprise, we observed similar trend of reduction in R_2_ values for both rostral-middle and caudal LC. Future studies are warranted for better delineation of rostro-caudal separation and whether other changes in tissue environments contribute to R_2_ values. Another notable observation is the strong relationship between LC integrity and verbal fluency. Several studies identify the impact of degraded LC integrity to lower verbal fluency [21, 22]. Interestingly, verbal fluency was more closely related to the integrity of the caudal LC, which innervates the spinal cord and cerebellum. While the primary function of the cerebellum is to coordinate voluntary movements, it is also consistently involved in language processing, and damage to the cerebellum can result in language disruption [84]. While further studies are needed to validate the underlying mechanisms, we speculate that degraded LC integrity contributes to compromised verbal fluency.

We also investigated the age-related differences in Myelin Water Fraction (MWF) derived from our BMC-mcDESPOT analysis [34]. MWF represents the fraction of the signal originating from water trapped within the myelin sheath, which experiences short T_1_ and T_2_ components. While a previous study revealed that LC neurons are sparsely myelinated [68], we found no significant age-related differences or associations with cognitive decline. This may be attributed to a technical limitation in our method for detecting low levels of MWF, or it could suggest that LC neurons do not exhibit measurable demyelination in healthy aging. Further research is needed to quantify and examine alterations in LC myelin.

There are limitations to the current study. First, participants were not recruited uniformly across all age intervals in our cohort. The number of participants between 50 and 65 years old was noticeably lower than other ages, which may have hindered the ability to observe potential inverted U-shape trajectories in our findings. While this might impact the interpretability of our findings in LC structural changes during normal aging, we obtained a meaningful sample size across the age range of our study. Furthermore, our cohort had a higher proportion of white participants, thus future studies with more diverse cohorts are warranted. Despite these limitations, our study has unique strengths. The extensive amount of longitudinal cognitive testing allowed for better characterization of longitudinal decline over long periods, yielding greater confidence in estimating rates of change. Secondly, our approach of registering to the MNI template may have obscured inter-subject variability in structural differences within the LC. However, since multiple studies demonstrated variability in LC identification [55], we aimed to minimize this inconsistency by utilizing a meta-mask that offers the highest agreement among all proposed masks. Future studies with higher spatial resolution and neuromelanin-sensitive imaging protocols can further address potential inter-subject variability in LC structures. Also, extending the investigation to other brain regions, such as the raphe nuclei, which are affected by tau pathology following LC degeneration, will further supplement our hypotheses and enrich our interpretation in how changes in LC may induce further deterioration in connected brain regions. Additionally, longitudinal imaging studies will provide more comprehensive understanding of how LC structural changes are evolving during aging process.

## 5. Conclusion

In this study, we explored the potential of quantitative MRI metrics to investigate age-related changes in the LC and their implication in cognitive dysfunction. Our results indicate that LC-qMRI relaxometry metrics are sensitive to age-related differences and cognitive decline. These quantitative biomarkers may offer a more accurate and reliable assessment of LC degeneration, enabling the monitoring of aging processes and facilitating the development of potential interventions to mitigate age-related diseases.

## Data Availability

The datasets generated and/or analyzed during this study are available upon reasonable request to the corresponding author, contingent upon institutional approval

## Declarations

- Ethics approval and consent to participate: Research protocols were approved by our institutional review boards and all participants were provided with written informed consents.
- Consent for publication: All participants provided their consents for publication.
- Availability of data and materials: The datasets generated and/or analyzed during this study are available upon reasonable request to the corresponding author, contingent upon institutional approval.
- Competing interests: All authors declare no conflicts of interest.
- Funding: This research was supported entirely by the intramural research Program of the NIH, National institute on Aging.
- Authors’ contributions: **Jonghyun Bae**: Conceptualization, Methodology, Formal analysis, Software, Writing-Original Draft, **Zhaoyuan Gong**: Visualization, Formal analysis, Writing-Review & Editing, **Caio Mazucanti**: Writing-Review & Editing, **Murat Bilgel**: Formal analysis, Data Curation, Writing – Review & Editing, **John Laporte**: Data Curation, Writing - Review & Editing, **Mary Faulkner**: Investigation, Writing - Review & Editing, **Alex Guo**: Software, Writing - Review & Editing, **Josephine Egan**: Writing - Review & Editing, Supervision, Funding acquisition, **Susan Resnick**: Writing - Review & Editing, Supervision, Resources, **Christopher Ramsden**: Writing - Review & Editing, Resources, Conceptualization, **Mustapha Bouhrara**: Conceptualization, Methodology, Writing – Original Draft, Writing - Review & Editing, Supervision
- Acknowledgements: Not applicable

## Notes

### Competing Interest Statement

The authors have declared no competing interest.

### Author Declarations

Ethics committee/IRB of National Institute on Aging gave ethical approval for this work.

